# Genomic epidemiology uncovers the timing and origin of the emergence of mpox in humans

**DOI:** 10.1101/2024.06.18.24309104

**Authors:** Edyth Parker, Ifeanyi F. Omah, Patrick Varilly, Andrew Magee, Akeemat Opeyemi Ayinla, Ayotunde E. Sijuwola, Muhammad I. Ahmed, Oludayo O. Ope-ewe, Olusola Akinola Ogunsanya, Alhaji Olono, Philomena Eromon, Christopher H Tomkins-Tinch, James Richard Otieno, Olusola Akanbi, Abiodun Egwuenu, Odianosen Ehiakhamen, Chimaobi Chukwu, Kabiru Suleiman, Afolabi Akinpelu, Adama Ahmad, Khadijah Isa Imam, Richard Ojedele, Victor Oripenaye, Kenneth Ikeata, Sophiyah Adelakun, Babatunde Olajumoke, Delia Doreen Djuicy, Loique Landry Messanga Essengue, Moïse Henri Moumbeket Yifomnjou, Mark Zeller, Karthik Gangavarapu, Áine O’Toole, Daniel J Park, Gerald Mboowa, Sofonias Kifle Tessema, Yenew Kebede Tebeje, Onikepe Folarin, Anise Happi, Philippe Lemey, Marc A Suchard, Kristian G. Andersen, Pardis Sabeti, Andrew Rambaut, Richard Njoum, Chikwe Ihekweazu, Idriss Jide, Ifedayo Adetifa, Christian T Happi

## Abstract

Five years before the 2022–2023 global mpox outbreak Nigeria reported its first cases in nearly 40 years, with the ongoing epidemic since driven by sustained human-to-human transmission. However, limited genomic data has left questions about the timing and origin of the mpox virus’ (MPXV) emergence. Here we generated 112 MPXV genomes from Nigeria from 2021-2023. We identify the closest zoonotic outgroup to the human epidemic in southern Nigeria, and estimate that the lineage transmitting from human-to-human emerged around July 2014, circulating cryptically until detected in September 2017. The epidemic originated in Southern Nigeria, particularly Rivers State, which also acted as a persistent and dominant source of viral dissemination to other states. We show that APOBEC3 activity increased MPXV’s evolutionary rate twenty-fold during human-to-human transmission. We also show how Delphy, a tool for near-real-time Bayesian phylogenetics, can aid rapid outbreak analytics. Our study sheds light on MPXV’s establishment in West Africa before the 2022–2023 global outbreak and highlights the need for improved pathogen surveillance and response.

---

Mpox is a viral zoonosis caused by the mpox virus (MPXV) of the genus *Orthopoxvirus*. MPXV diversity is partitioned into two major clades: Clade 1, which is endemic in an as-yet unknown non-human animal reservoir in Central Africa, and Clade II with subclades Clade IIa and Clade IIb, similarly endemic in Western Africa.^1,2^ From the first recognized human case in 1970 in the Democratic Republic of Congo (DRC) until 2017 there have only been occasional outbreaks and sporadic cases with limited human-to-human transmission in endemic regions.^3–5^

In September 2017, Nigeria reported its first mpox outbreak in nearly 40 years, with a larger second wave of cases towards 2022.^5,6^ The outbreak had distinct epidemiological characteristics not typically seen with MPXV zoonotic infections. There was a marked demographic shift to more urban, adult individuals of 30 to 40 years of age, compared to more typical cases reported in children in rural communities.^6,7^

In May 2022, approximately five years after the 2017 outbreak in Nigeria, a Clade IIb lineage termed B.1 rapidly disseminated around the world to cause the global mpox outbreak.^8^ The apparent human-to-human transmission of the B.1 lineage raised the possibility of a new transmission route for MPXV. B.1 showed significant divergence from the closest Clade IIb genome sampled in Nigeria in 2018, with an evolutionary rate significantly elevated above the expected rate for Orthopoxviruses.^9^ The global outbreak was characterised by enrichment of mutations in a specific dinucleotide context (TC→TT or the reverse complement GA→AA) associated with the cytosine deaminase activity of the APOBEC3 (apolipoprotein B mRNA editing enzyme, catalytic polypeptide 3) host antiviral mechanism.^10,11^ This mutational signature has not been observed in sequences from zoonotic infections, strongly suggesting that APOBEC3 genomic editing was a characteristic feature of sustained transmission in the human population.^11^

In light of this new evolutionary dynamic, several studies confirmed that the ongoing mpox epidemic in Nigeria was driven by sustained human-to-human transmission, not independent zoonotic infections or self-limiting transmission chains.^11,12^ It is estimated that MPXV emerged in the human population in Nigeria in 2016, circulating and diversifying cryptically into multiple distinct lineages.^11,12^ The diversity circulating during the human outbreak in Nigeria, from which the B.1 global lineage descended, is referred to as hMPXV-1, as designated by Happi *et al*.^13^

However, the limited availability of high-quality genomes and their restricted geographic representation leaves several uncertainties around the transmission dynamics of hMPXV-1 during the ongoing human epidemic in Nigeria. The genomic data supports a single zoonotic origin for hMPXV-1.^11^ However, no close zoonotic outgroup has been identified to date, and estimates of the timing of hMPXV-1’s emergence in the human population is based on limited genomic data. The evolutionary dynamics of hMPXV-1 after the initial spillover remains largely unknown, with a limited genomic picture already revealing multiple distinct lineages co-circulating.^12^ Importantly, it is not known whether all cases in Nigeria are part of the human epidemic, or whether there are ongoing spillover events resulting in independent onward transmission. It is also not clear what the drivers of this sustained human transmission are, and whether there are clear spatiotemporal spread patterns seeding and reseeding local epidemics across Nigerian states.

To address the open questions around the origin, spread, and evolution of the recent mpox outbreak, we formed a Pan-African consortium to collate the largest MPXV dataset from Africa to date. In this study, we generated 112 genomes sampled from Nigeria from 2021 to 2023, including the period before the emergence of the B.1 global outbreak. We re-estimate the date of MPXV’s transition to sustained human-to-human transmission using a custom phylodynamic model. With the widest geographic representation sampled in Nigeria yet, we reconstruct the spatiotemporal spread of hMPXV-1 across Nigeria and quantify the drivers of hMPXV-1 transmission.

## Results

### Distinct hMPXV-1 lineages co-circulate during the ongoing human epidemic in Nigeria

To investigate whether all of our mpox samples were part of the human epidemic, as well as the extent of hMPXV-1’s cryptic diversification across the epidemic, we generated 112 MPXV genomes sampled across Nigeria from July 2021 to May 2023. The median coverage of the sequences generated ranged from 30 to 7900-x. Our sequences were predominantly sampled from the South South, South East, and South West regions of Nigeria (Figure 1 A, B). The South region was the epicentre of the epidemic from 2017 onwards, with southern states reporting the earliest cases (Figure 1 A, B). From 2022 onwards, both the northern and southern regions experienced a substantial resurgence of cases after a period of low incidence. Our dataset largely encompasses the resurgence from 2022 onwards (Figure 1A). All of our sequences belonged to Clade IIb in the global phylogeny (Figure 1C).

To investigate hMPXV-1’s cryptic diversification with our expanded dataset, we reconstructed the Clade IIb phylogeny with our 105 non-B.1 novel genomes and all available Clade IIb sequences.^12^ Within hMPXV-1, distinct lineages are designated according to a system similar to the SARS-CoV-2 Pango nomenclature to track the lineages circulating within the outbreak.^13,14^ Under this nomenclature hMPXV-1 is referred to as Lineage A, with its descendants designated as e.g. A.1 and A.2 and second subdivision descendants designated as e.g. A.1.1. We found that 105 of our sequences were interspersed throughout six divergent, co-circulating sub-lineages of hMPXV-1 (Lineage A) (Figure 1D, E). Lineage A represents the sustained human-to-human transmission reported in Nigeria from 2017 onwards, as well as travel-associated cases and the B.1 global outbreak.^11–13^ The majority of our sequences belonged to sub-lineage A.2.3 (Figure 1E). We did not find any geographic structure in hMPXV-1, indicating that the sub-lineages were not confined to specific geographic subpopulations. We also identified three B.1 sequences, representing re-introductions from the global epidemic into Nigeria. Notably, none of our sequences were from lineage A.1.1, from which the B.1 global outbreak lineage descended (Figure 1D).

We investigated whether all of our sequences formed part of the ongoing human epidemic or whether any of them represented zoonotic transmission events by quantifying the APOBEC3 mutational bias characteristic of human-to-human transmission across our phylogeny.^11,15,16^ We performed ancestral state reconstruction across our Clade IIb phylogeny to map single nucleotide polymorphism (SNPs) to their relevant branches. We annotated APOBEC3 characteristic mutations i.e. C→T or G→A in the correct dimer context along branches (Figure 1D) and calculated their relative proportion across internal branches (Figure 1F).

We observed a significant proportion of mutations consistent with APOBEC3 activity across the internal and terminal branches of hMPXV-1, including our sequences (Figure 1D, F). Approximately 74% of reconstructed SNPs in hMPXV-1 were indicative of APOBEC3 editing, consistent with previous work.^11^ In contrast, only 9% of reconstructed SNPs across the zoonotic parts of the tree (excluding the subtree annotated in blue) were APOBEC3-type mutations (Figure 1F). This APOBEC3 enrichment indicates that 105 of our 109 non-B.1 sequences form part of the sustained human-to-human transmission underlying the Nigerian epidemic. Notably, two sequences did not show APOBEC3 enrichment, indicating these represent zoonotic transmission events (outgroup “Zx” in Figure 1D). Taken together, these findings suggest that hMPXV-1 has cryptically circulated and diversified, largely driven by APOBEC3 activity, in the human population without geographic restrictions in Nigeria for a prolonged period after initial spillover.^11,12^

**Figure 1:**
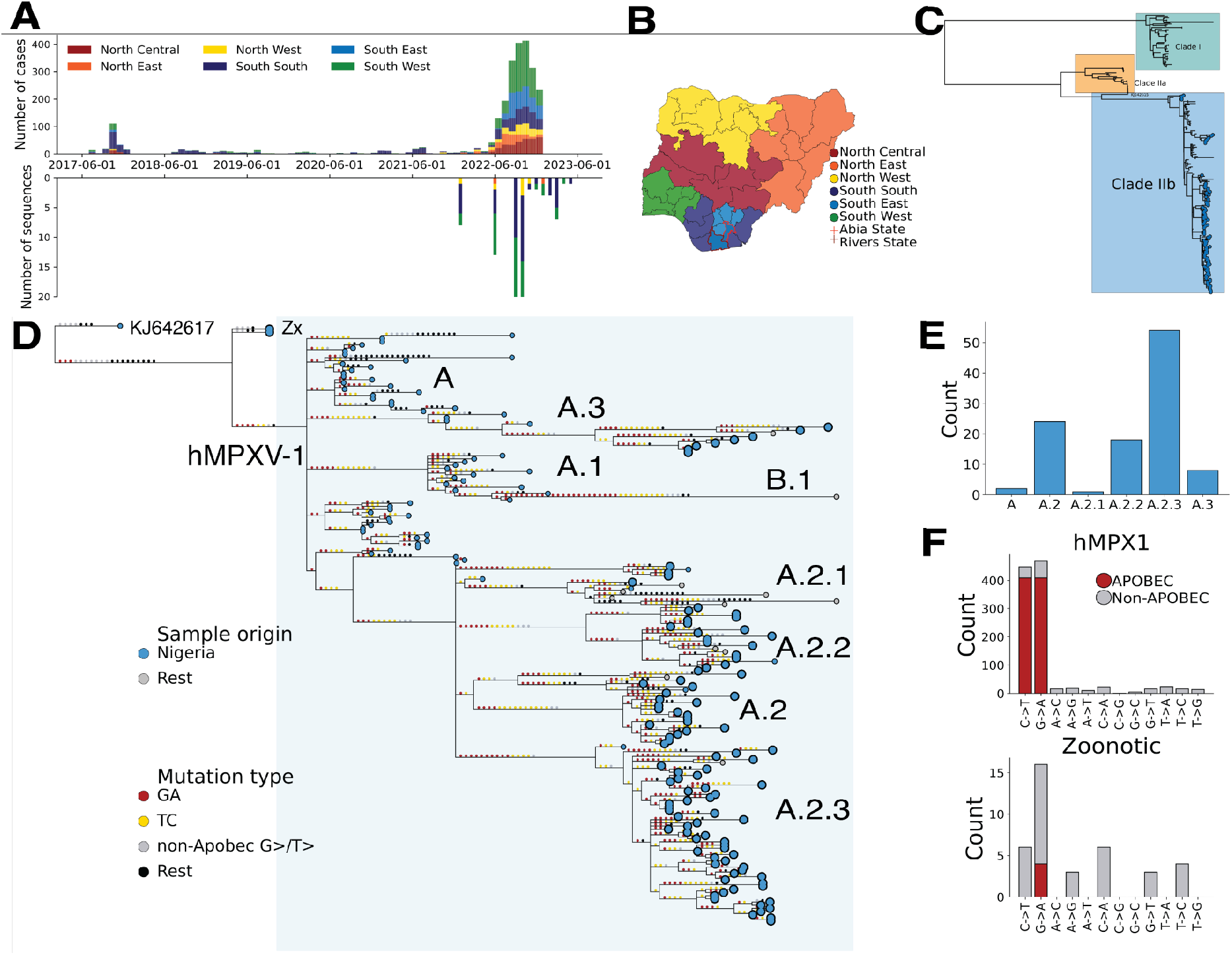
**A**) Epidemiological incidence of mpox cases in Nigeria coloured by geopolitical region (top panel), relative to our genomic dataset’s temporal and geographic distribution (bottom panel). **B)** Geopolitical regions of Nigeria, with Abia and Rivers State highlighted with red borders **C)** Global MPXV phylogeny of Clade I, Clade IIa and Clade IIb. Our Clade IIb sequences are annotated with tip points **D)** Clade IIb phylogeny with reconstructed SNPs mapped onto branches. APOBEC3 mutations along the branches are annotated in yellow and red, with the remainder in grey and black. The hMPXV-1 Clade (Lineage A) is annotated and highlighted in the light blue box, with the lineage annotation in text. Our new zoonotic outgroup sequences are annotated as “Zx”. Our sequences are highlighted as enlarged tips relative to the background tips. **E)** Lineage distribution of our Clade IIb sequences. **F)** The number of overall reconstructed SNPs that APOBEC3 substitutions accounts for in the hMPXV-1 subtree (highlighted and annotated in blue in Figure D) and the Zoonotic outgroup (KJ642617 and Zx annotated in Figure D).

### Closest zoonotic outgroup to hMPXV-1 circulated in southern Nigeria

We explored whether the two zoonotic sequences we identified that lack APOBEC3 signal might represent a closer zoonotic outgroup to hMPXV-1 than the current outgroup (KJ642617), sampled in 1971 (Figure 1D). We found that they form a sister lineage to hMPXV-1 (“Zx’’ in Figure 1D), breaking up the long stem branch from KJ642617 to hMPXV-1. As our new sequences share a common ancestor with hMPXV-1, they represent the closest zoonotic outgroup to hMPXV-1 (Figure 1D).^17^ The Zx sequences reduced the branch from a zoonotic outgroup to hMPXV-1 from 27 to 8 SNPs. Our new Zx sequences were sampled in Abia State in southern Nigeria, where KJ642617 was also sampled in 1971. Abia State provides suitable ecological conditions to host reservoir populations where zoonotic precursors may have circulated.^17^

A previous study found that the genomic data supported a single zoonotic origin for hMPXV-1, as all of the sequences from the human epidemic shared a substantial number of APOBEC3 mutations that occurred along the stem branch from KJ642617 and hMPXV-1’s zoonotic common ancestor.^11^ With our new zoonotic outgroup, we investigated the number of shared APOBEC3 mutations along the stem branch. We found that all of the diversified hMPXV-1 sub-lineages shared six APOBEC3 mutations (Figure 1D). This supports findings that it is unlikely that the emergence of hMPXV-1 in the human population resulted from repeated independent zoonotic events.^11^

### The mpox epidemic has grown exponentially, but slowly, since emergence in 2014

Our new zoonotic outgroup breaks up the long stem branch from hMPXV-1’s most recent common ancestor (MRCA) in non-human animals. This more recent divergence of our zoonotic outgroup and hMPXV-1 places a tighter bound on when hMPXV-1 could have emerged in the human population. To estimate the timing of MPXV’s initial emergence in the human population with our newly identified zoonotic outgroup and our expanded dataset, we adopted the two-epoch model of O’Toole *et al.* implemented in the BEAST software package.^11^ The model explicitly accounts for APOBEC3-mediated evolution by allowing for the transition from a background evolutionary rate driven by the polymerase error rate to an APOBEC3 driven rate across the tree in a partitioned alignment (see Methods).

We estimated that the transition to sustained human transmission, representing the time of emergence in the human population, occurred in mid July 2014 (95% HPD 6 October 2013 - 23 February 2015) (Figure 2A). Our estimate is approximately 14 months earlier than previous reports, though the credible intervals overlap (vs 14 September 2015, 95% HPD 21 August 2014 to 31 July 2016).^11^ We estimated that time to the most recent common ancestor or tMRCA of hMPXV-1 was mid July 2015 (median time to the most recent common ancestor or tMRCA, 95% HPD 8 November 2014 - 22 February 2016), representing the time at which hMPXV-1 started to diversify in humans (Figure 2A). Our estimate is approximately eight months earlier than previous, though credible intervals overlap (vs 23 February 2016, 95% HPD 28 June 2015, 28 September 2016).^11^ Collectively, our results indicate that hMPXV-1 circulated cryptically in the human population in Nigeria for approximately three years before detection during the late 2017 wave of cases, and more than seven years before disseminating globally during the B.1 outbreak in 2022. This prolonged period of cryptic human-to-human transmission likely explains the diversification of hMPXV-1 into the distinct sub-lineages we observed (Figure 1D).

After the initial surge of cases in 2017 there was a period of low reported incidence between 2018-2022, including the early years of the COVID-19 pandemic (Figure 1A). This low incidence, especially from 2020-2021, may be attributable to underreporting associated with the impact of COVID-19 on health systems, as well as COVID-19 related public health interventions. To investigate whether the mpox epidemic had grown continuously since emergence or whether there was a real decline in cases during the period of low reported incidence, we allowed for a two-phase coalescent model in our epoch-model. Under the model, the tree from the MRCA(hMPXV-1) onwards was modelled with an exponential growth model, and the earlier zoonotic phases were modelled under a constant-population size model. We also estimated the population dynamics of hMPXV-1 alone under a non-parametric Skygrid model. We found evidence of exponential growth following hMPXV-1’s emergence under both models (Figure 2B), as previously reported.^11^ This indicates that the epidemic was growing exponentially even during periods of low reported incidence (Figure 1A) with an estimated doubling time of approximately two and a half years (Figure 2C). This exponential, but slow growth rate suggests that the epidemic has not spread to the general population, but is concentrated in a more restricted subpopulation.^18^

**Figure 2.**
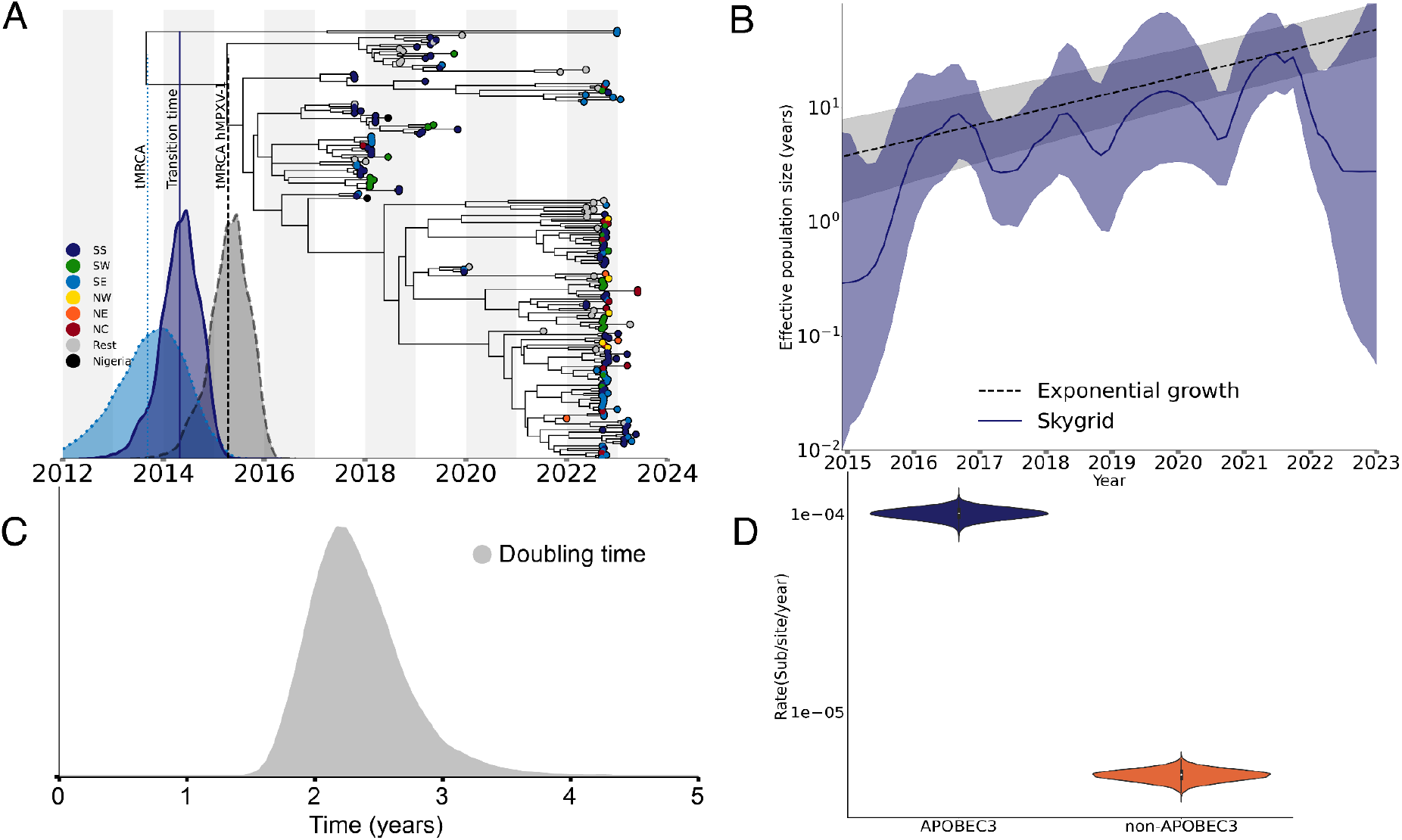
**A)**. Bayesian maximum clade credibility (MCC) tree of Clade IIb indicating the time of emergence of hMPXV-1 into the human population. The distributions indicate the 95% HPD for 1) the tMRCA of the closest zoonotic outgroup of the hMPXV-1 2) the time of transition to sustained human-to-human transmission and 3) the tMRCA of hMPXV-1. SS: South South; SW: South West; SE: South East; NW: North West; NE: North East’ NC: North Central. **B**) The effective population size of the epidemic in Nigeria under a Skygrid and exponential coalescent model **C**) The posterior distribution of the estimated doubling time of the epidemic since emergence **D**) Estimates of the APOBEC3 and Non-APOBEC3 clock rates.

### APOBEC3 activity increased the evolutionary rate 20-fold during sustained human transmission

APOBEC3-mediated genomic editing has significantly elevated the evolutionary rate of hMPXV-1 above the expected rate for double-stranded DNA viruses.^11^ To investigate how much higher the APOBEC3-mediated evolutionary rate is relative to the polymerase-driven rate in the context of human-to-human transmission, we employed the partitioned two-epoch model. We estimated an APOBEC3 clock rate of 1×10^-4^ substitutions per site per year (subs/site/year) (95% HPD 8.8×10^-5^ - 1.14×10^-4^), and a background evolutionary rate of 4.8×10^-6^ subs/site/year (95% HPD 4.2×10^-6^ - 5.5×10^-6^) (Figure 2D). This indicates that APOBEC3 activity increased the rate ∼20 times higher relative to the background evolutionary rate during the human epidemic in Nigeria.

### Southern Nigeria was an early and persistent source of hMPXV-1 dissemination from emergence onwards

Nigeria’s southern states were the early epicentre for the mpox epidemic, and reported the highest number of cases throughout (Figure 1A). Despite the clustering of the earliest cases in South South states, the geographic origin of hMPXV-1 remains unknown. The northern states only reported a significant number of cases after the resurgence in 2022 (Figure 1A). However, it is not known whether there was widespread, under-ascertained and unsampled transmission outside of the southern states before the resurgence in 2022. It is also not clear which states contributed to interstate viral dissemination and how these patterns may have shifted across the different epidemic phases. To better understand the spatiotemporal spread of hMPXV-1 within Nigeria, we used discrete and continuous phylogeographic reconstructions on a state and regional level. We found that both the reconstructions support that hMPXV-1 likely originated in Rivers State in the South South (Figure 3A, Posterior = 0.91, Extended Data Figure 1). The MRCA of hMPXV-1 circulated in Rivers State in July 2015 (tMRCA, 95% HPD 8 November 2014 - 22 February 2016) (Figure 3A). Notably, this is consistent with our sampling of the closest zoonotic outgroup to hMPXV-1 in the neighbouring southern Abia state (Figure 1D).

We found that Rivers State was the primary source of interstate viral exports across the epidemic, with an estimated 72 introductions originating in Rivers (95% HPD: 62-80) (Figure 3 A, B, Extended Data Figure 1). The highest number of viral exports from Rivers spread to other South South states as well as the South West, followed by the South East (Figure 3B, Extended Data Figure 2). Overall, neighbouring Imo and Lagos in the South West had the highest number of introductions from Rivers, followed by neighbouring Bayelsa. The remainder of the South South states as well as the South East and South West states were all equivalently the second highest source of viral exports overall.

We found that all introductions in the early epidemic originated in Rivers State (Figure 3C). Viral spread from Rivers into neighbouring South South states such as Bayelsa and Imo, as well as Lagos in the South West, occurred as early as 2016 (Figure 3C, Extended Data Figure 3). This is consistent with the epidemiological data that confirms southern Nigeria as the epicentre of the early epidemic, with the first case reported in Bayelsa on 11 September 2017 (Figure 1A). All save one sampled introductions into Northern states occurred in the later phase of the epidemic, after the resurgence towards 2022 (Figure 3C, Figure 1A). This spatiotemporal pattern was consistent across our discrete and continuous phylogeographic reconstructions on both a regional and state level (Figure 3D, Extended Data Figures 1-3).

To investigate how widespread early hMPXV-1 transmission was across Nigeria by the time the outbreak was declared on 22 September 2017, we performed a continuous phylogeographic reconstruction. We found that the virus had spread more than 500 km beyond Rivers State into Bayelsa, Imo, Delta, Edo, FCT, and Lagos before the first case was detected on 11 September 2017 (Figure 3D). Collectively, this suggests that the human epidemic originated in Rivers State, with early spread of the virus to neighbouring South South and South East states and Lagos before outbreak declaration, and with delayed dispersal to the north.

**Figure 3:**
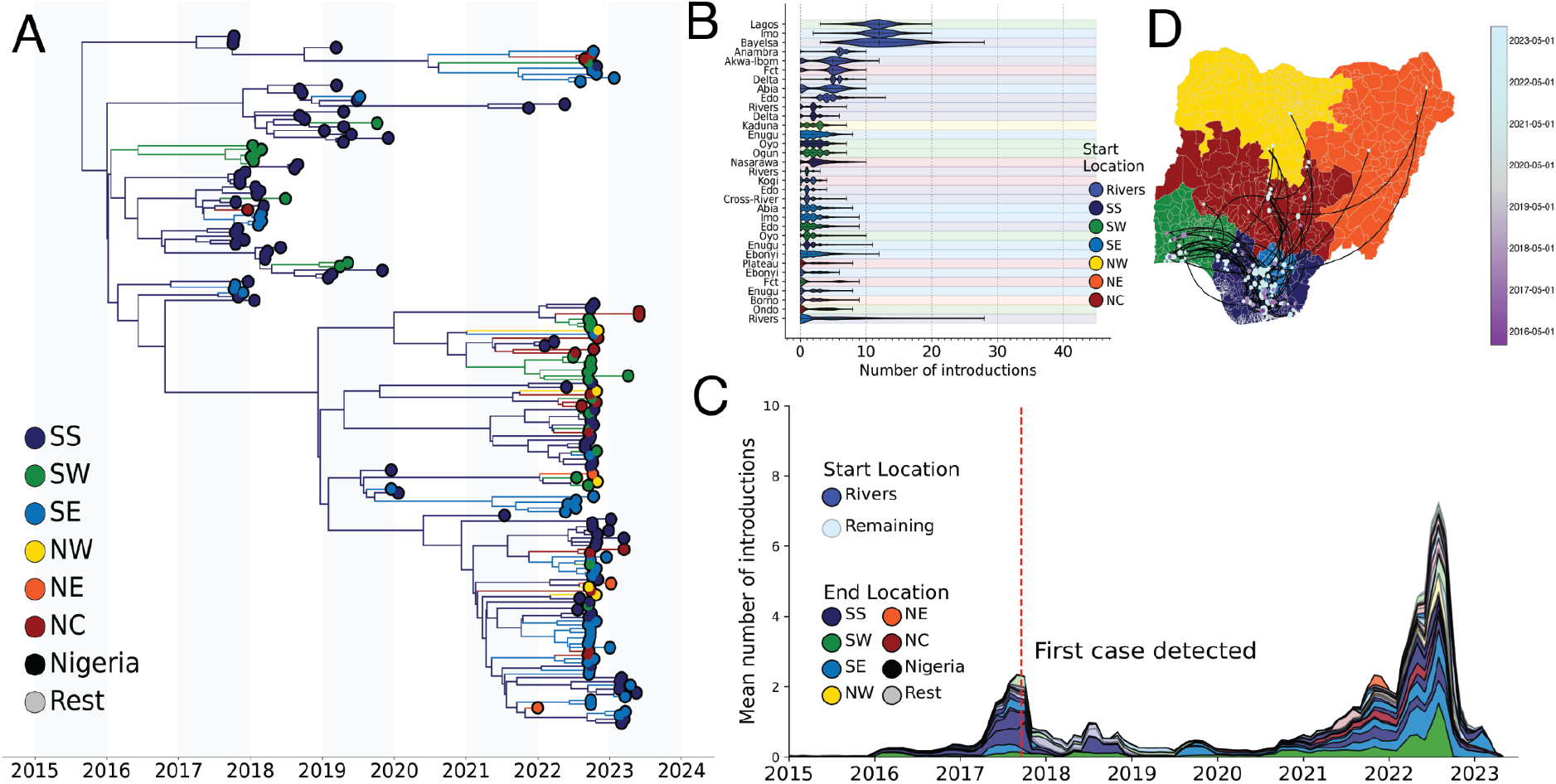
**A)** Phylogeographic reconstruction of the spatiotemporal spread of Clade IIb in Nigeria. The branches of the Maximum Clade Credibility tree (MCC) are coloured by source region, as per legend. SS: South South; SW: South West; SE: South East; NW: North West; NE: North East’ NC: North Central. **B)** Total number of introductions by state from each start location, annotated as per legend in colour, to each end location on the y-axis. The regions of the end location state on the y-axis are highlighted in colour in the plot background as per the legend. **C)** The distribution of the number of introductions across time by state. The end location state is coloured by region, as per legend. The start location is highlighted by transparency: all introductions originating from Rivers State are presented with no transparency, whereas introductions originating from other states are more transparent. **D)** Continuous phylogeography of hMPXV-1 spatiotemporal spread across Nigeria, with timing of viral dissemination highlighted by colour range as per legend.

It is not clear whether cases across Nigerian states were continuously seeded by interstate introductions from Rivers and the south, or whether there were locally persistent transmission chains in other regions sustaining local epidemics. Towards understanding the respective contribution of persistence and introductions, we investigated the persistence of transmission chains in each state. We found that hMPXV-1 has persistently circulated in Rivers from emergence onwards (Figure 4A, B). hMPXV-1 diversified in Rivers State for more than two years before the first case was reported in Bayelsa on 11 September 2017 (Figure 4A, Extended Data Figure 4).^5,6^ When the outbreak was declared, transmission chains had already been established in 15 states outside of Rivers (Figure 4A). Delta and Bayelsa in the South South had the second longest persistence of a transmission chain at approximately four and a half and three years respectively, followed by the earliest chain established in Lagos at three years (Figure 4 A,B). Outside of Rivers State and the early chains in Bayelsa and Lagos, the longest persistence was estimated for lineages introduced during the period of low reported incidence in 2018-2021, when sampling was sparse (Figure 4B, 1A).^19^

We found that persistent transmission was the primary driver of the epidemic in Rivers State, relative to repeat introductions (Figure 4A, C). However, the percentage of transmission chains persistently circulating was dynamic over time in other South South states (Figure 4 D). Transmission chains seeded by Rivers State early in the epidemic in the South South, South West and South East only persisted locally for less than two years, excluding the first transmission chains established in Bayelsa and Lagos (Figure 4 A, B). There was a significant increase in the number of transmission chains circulating during the resurgence of cases towards 2022 (Figure 4C). This is consistent with the increased viral exports during this period (Figure 3C), seeding new transmission chains that drove local surges across Nigeria (Figure 1A, Figure 4A). Transmission chains from the later stage of the epidemic were predominantly introduced from Rivers, and persisted for less than two years (Figure 4A). However, it is unclear whether this pattern persists past the end of our sampling frame. There was no evidence for significant persistence in Northern states prior to the later phase of the epidemic, which is consistent with the low reported incidence and delayed viral spread observed (Figure 4 A, 3D, FIgure 1A). Altogether, this further supports that Rivers State acted as the persistent source for the epidemic, while local epidemics in other states were largely driven by repeat introductions.

To account for uneven sampling across states, we also performed our phylogeographic analyses at the regional level. We observed a consistent pattern to our state-level analyses: early and predominant spread from the South South, with initial spread to the South East and South West. There was strong evidence of persistent circulation in the South South, with local epidemics in all other regions driven by repeated introduction from the South South (Extended Data figures 1–4).

**Figure 4:**
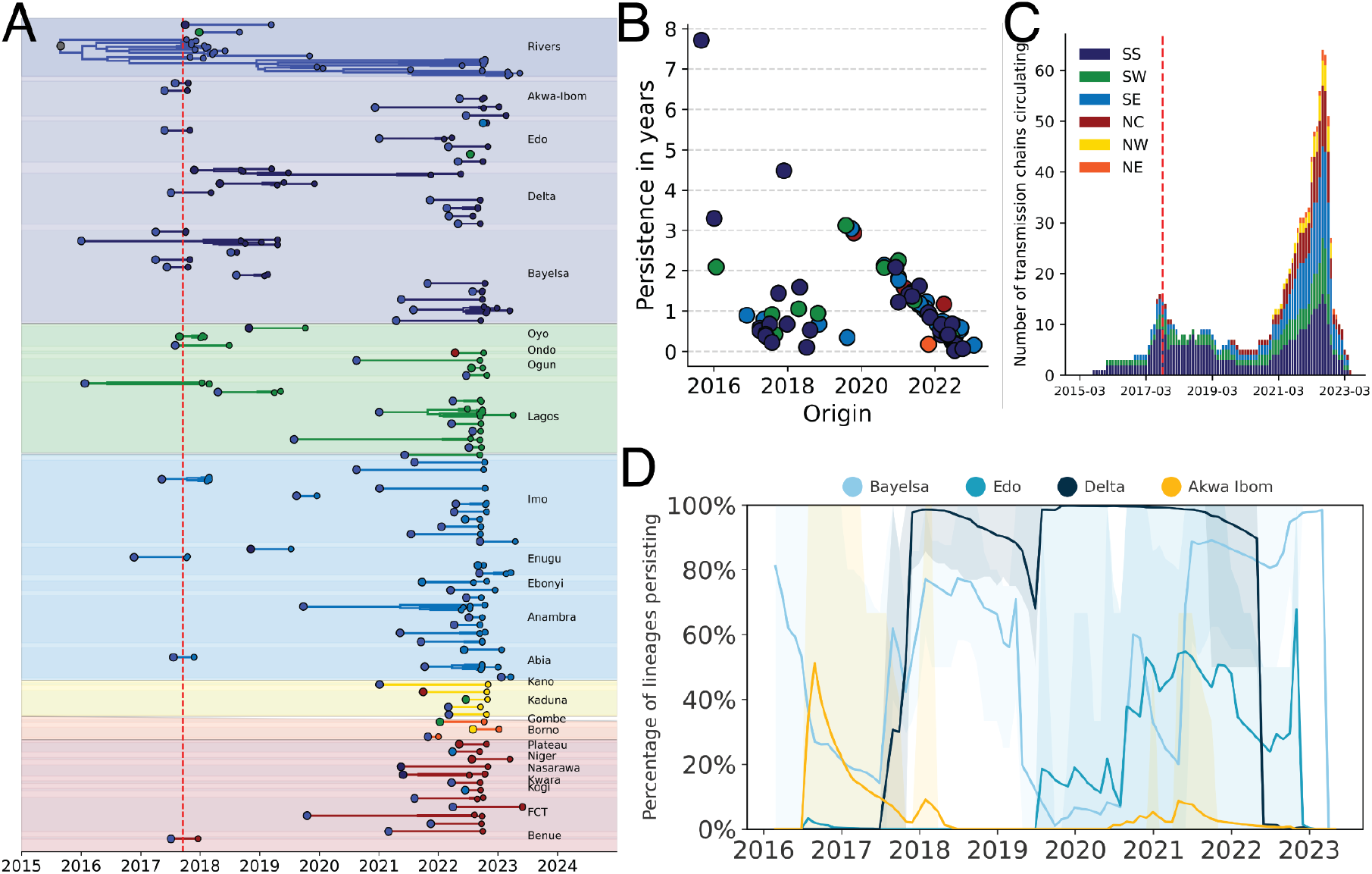
**A)** Persistence of transmission chains across all Nigerian states sampled. Individual chains are coloured by region, with the boundary of each individual state highlighted by a filled background and annotated in text on the right. The start of each transmission chain is coloured by its state of origin. The red line indicates the date of report for the first case in Bayelsa on 17 September 2017. **B)** The persistence in years of each transmission chain across its time of origin, coloured by region. **C)** The number of transmission chains circulating across all regions across time, calculated by a month-sliding window. The red line indicates the date the outbreak was declared on 22 September 2017. SS: South South; SW: South West; SE: South East; NW: North West; NE: North East’ NC: North Central. **D)** The percentage of transmission chains persisting across time for South South states excluding Rivers.

### Rivers State origin was associated with hMPXV-1 dispersal

Our phylogeographic reconstructions consistently support a spatiotemporal pattern of early viral spread between and then from southern states, with hMPXV-1 persistently circulating in the South South region. To identify potential drivers of this pattern, we used a phylogeographic generalised linear model that integrates covariates of spatial spread to determine what factors were associated with hMPXV-1 dispersal. We incorporated covariates in our model including epidemiological, demographic, geographic and economic variables, as well as location-specific predictors that capture a pairwise binary transition across different states (see Extended Data table 1).

Of the 19 covariates analysed, we found that the main predictor that positively affected viral dispersal was whether the lineage originated in Rivers State (BF>50) (Figure 5A, B). There was no support for the residual covariates that assessed the deviations of sampling numbers relative to epidemiological cases (Figure 5A). This suggests that the out-of-Rivers pattern is robust and was not due to sampling heterogeneity across locations. This finding supports our previous analyses highlighting Rivers’ early and dominant role in the spread of hMPXV-1 from emergence onwards (Figure 5B). The population density in the destination was also positively associated with hMPXV-1 dispersal (BF>15), which is consistent with the principles of a gravity model in epidemiology (Figure 5A).^20,21^

**Figure 5:**
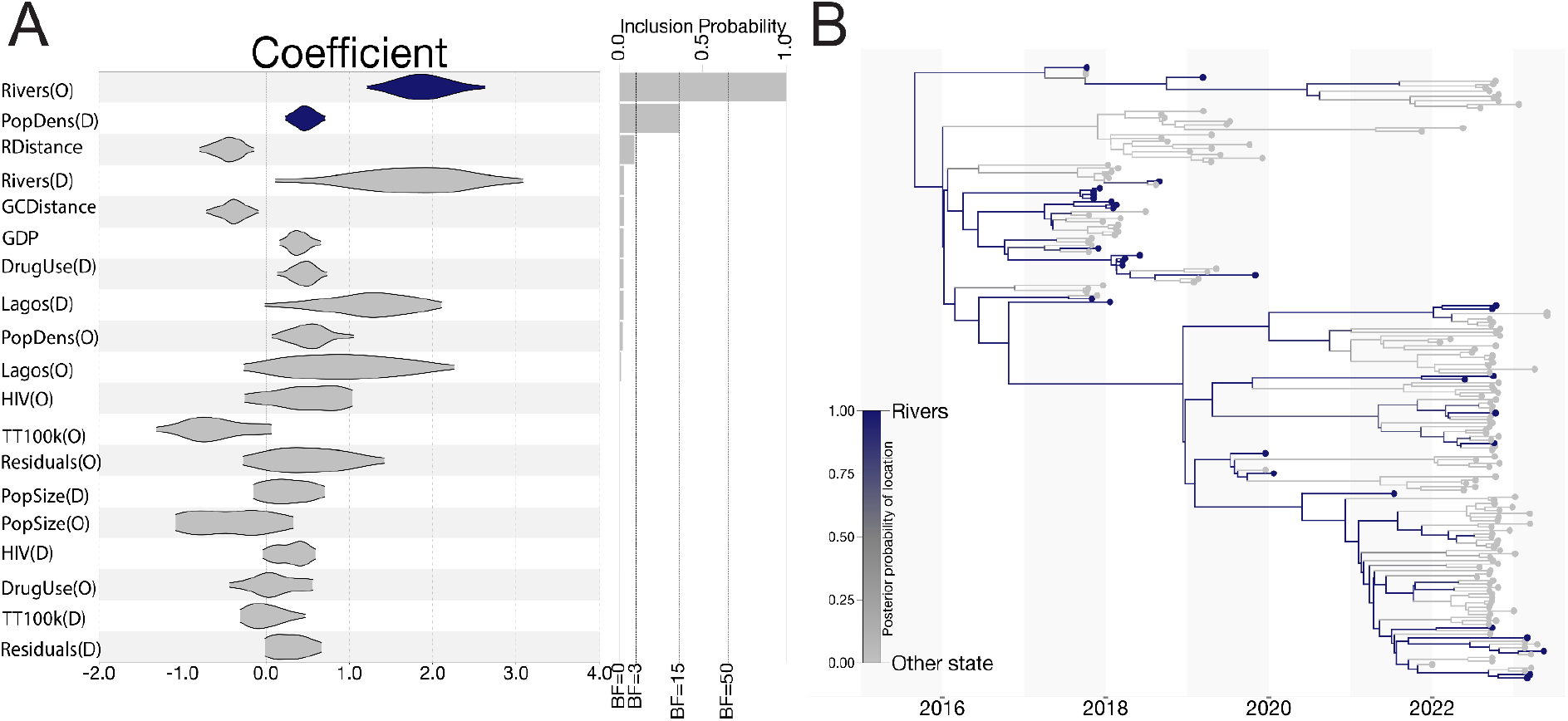
**A)** The GLM coefficients of covariates of spatial spread and their associated Bayes Factors. Significant covariates are highlighted in purple. **B)** Phylogeographic reconstruction of the migratory pattern of hMPXV-1 from Rivers to other Nigerian states.

### Informing real-time public health decision making with Delphy

Effective outbreak responses rely on rapid action and real-time information to guide decision making. To provide public health professionals with a broadly accessible tool for near-real-time Bayesian phylogenetics, we have developed a new tool named Delphy.^22^ Central to Delphy’s speed is a reframing of Bayesian phylogenetics that exploits the characteristics of typical genomic epidemiology datasets to make it both faster and more scalable (see ^22^, Methods and SI). Delphy is accessible as a client-side web application at https://delphy.fathom.info, which provides an intuitive interface for analysing the entire posterior distribution of trees.^23^

To demonstrate Delphy’s utility in outbreak response analytics, we adapted it to analyse our mpox sequences from the human epidemic in Nigeria. For this analysis, we assume that any zoonotic spillover event precedes the MRCA of hMPXV-1 (see Methods). We found that Delphy produced consistent estimates with our analogous BEAST runs, with credible intervals overlapping (Figure 6A, 2A). The tMRCA estimate of the hMPXV-1 clade from Delphy was 18 Jan 2016 (95% HPD 17 Jun 2015 to 1 July 2016), compared to the BEAST estimate of 15 July 2015 (95% HPD 8 November 2014 - 22 February 2016). The estimates for the APOBEC and polymerase-driven substitution rate as well as the doubling time of hMPXV-1 were also consistent: for the polymerase, it was 8.6×10^-6^ substitutions per site per year (95% HPD 7.4×10^-6^-9.9×10^-6^) with Delphy and 9.7×10^-6^ substitutions per site per year (95% HPD 8.2×10^-6^-1.1×10^-5^) for BEAST; for the APOBEC-driven substitution rate the estimates were 1.1×10^-4^ substitutions per site per year (95% HPD 1×10^-4^-1.3×10^-4^) for Delphy and 1×10^-4^ substitutions per site per year (95% HPD 8.8×10^-5^ - 1.14×10^-4^) for BEAST; the doubling time estimates were 2 years (95% HPD 1.6 - 2.5 years) for Delphy and 2.2 years (95% HPD 1.7 - 2.9) for BEAST.

Delphy can also be used to reconstruct simple estimates of tip-associated metadata at the inner nodes of a Maximum Clade Credibility tree (MCC) with a simple parsimony algorithm. We found that the simplified parsimony approach recovered spatial dynamics consistent with the phylogeographic patterns reconstructed in the BEAST analyses (Figure 6A vs 1D, Figure 6B vs Figure 3B, Extended Data Figure 5).

Delphy allows public health professionals to approximate the type of outbreak analytics demonstrated in this paper in an accessible client-side web application in near real-time. The Delphy run was completed in 30 minutes on a standard 4-core 2020 laptop without a GPU, relative to the 8 hour run time of the BEAST analyses on a standard M1 Macstudio.

**Figure 6:**
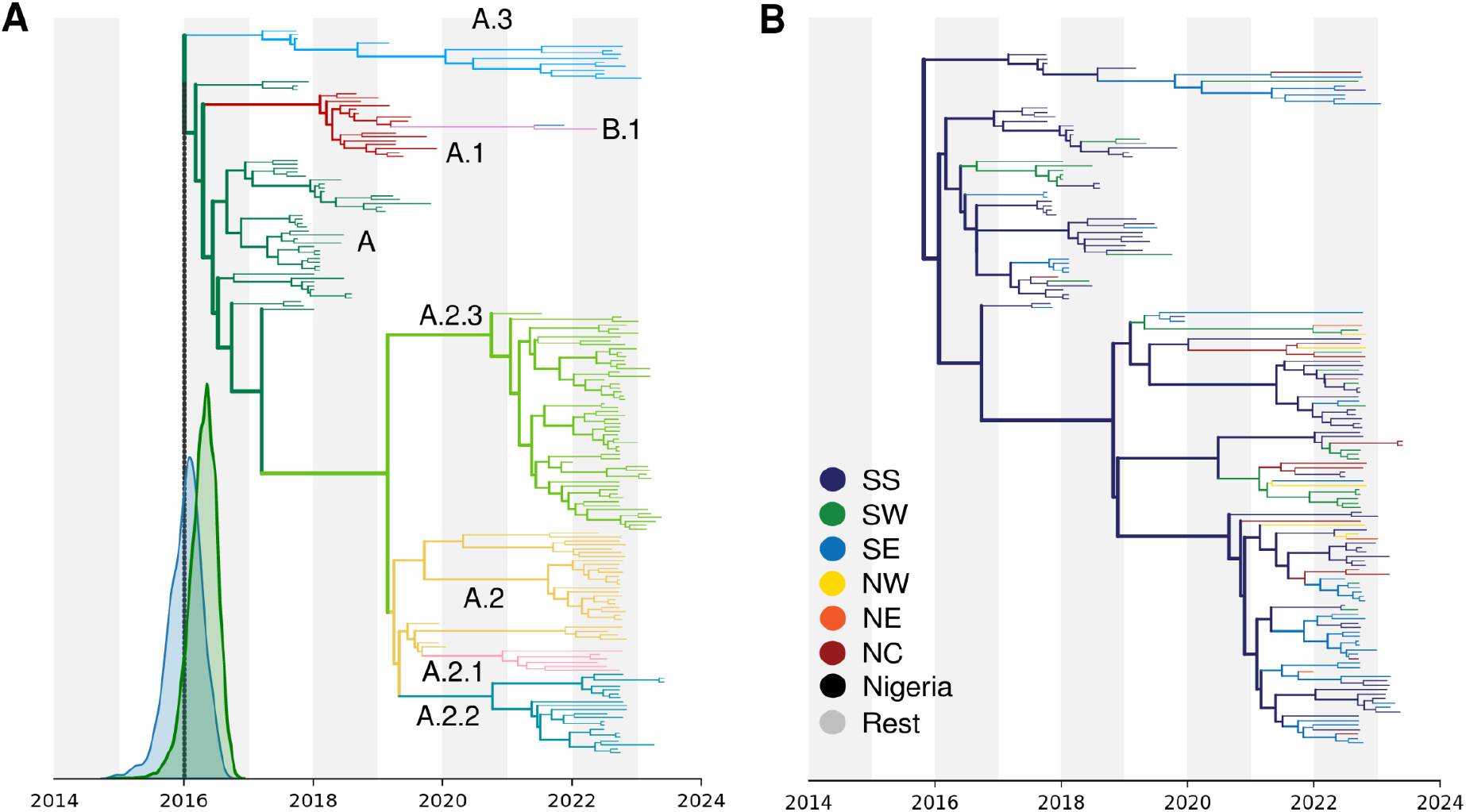
**A)** Delphy MCC for hMPXV-1 samples, coloured by nomenclature^13^ (hosted at https://delphy.fathom.info/?mpox2024-post-spillover-with-metadata.dphy); compare to Figs 2A and 1D. The tMRCA distribution calculated by Delphy (blue) overlaps with that of an analogous BEAST run on the same sequences (green); the run for Figure 2A also includes the remaining zoonotic sequences. **B)** Delphy MCC for hMPXV-1 sequences from Nigeria only (hosted at https://delphy.fathom.info/?mpox2024-post-spillover-nigeria-only-final-with-metadata.dphy), coloured by geographical origin at the tips and propagated to inner nodes via naive parsimony; compare to Fig 3A.

## Discussion

Consensus evidence now strongly supports that MPXV Clade IIb is no longer solely a zoonotic disease but is now sustained in a human subpopulation in Nigeria, where it has been circulating cryptically for nearly a decade. However, the drivers behind this emergence in humans remains uncertain.^17^ It is not clear why this cross-species transmission event did not simply result in a sporadic case or a self-limiting transmission chain as all previous zoonotic MPXV infections. Nonetheless, it is likely that the spillover event occurred in a more connected, mobile subpopulation in Nigeria with more probable onward transmission driven by behavioural or demographic factors.^17^

The epidemic in Nigeria has already resulted in the global B.1 outbreak, as well as several self-limiting exports of distinct Clade IIb sub-lineages detected in countries with higher levels of surveillance.^16^ It is possible that there is related or independent cryptic sustained human-to-human transmission in other regional countries with less robust surveillance systems, including for Clade IIa and Clade I.^24^ However, we did not detect any evidence of sustained human-to-human transmission in Cameroon.^17^ It is highly likely that viral export and potential seeding of outbreaks, both regional and intercontinental, will continue as the virus circulates in the human population. It is also possible that sustained circulation will result in viral adaptation through APOBEC3-accelerated evolution across larger chains of infection.^25^ It is therefore vital that the Nigerian epidemic is resolved or reduced by targeted public health interventions. Our study has provided critical insights to facilitate strategic public health interventions. Across our phylogeographic reconstructions, we found that Rivers State and other South South states served as early, dominant and persistent sources of viral export. Interventions should accordingly be targeted to these regions. Notably, we find evidence of prolonged cryptic circulation and geographic expansion before detection in these regions, emphasising the need for enhanced surveillance and improved diagnostic and surveillance infrastructure in these regions. We also estimated a relatively slow doubling rate, suggesting that the virus is circulating in a restricted subpopulation.^25^ Enhanced surveillance of cases is required to characterise the underlying transmission network and associated risk factors, allowing for targeted interventions before the epidemic becomes more generalised.

However, our results should be interpreted within the limits of our sample. Our sample represents all available and viable samples from routine surveillance, but were not systematically selected for spatiotemporal representativeness. However, our sample represents 4.2% of all suspected mpox cases in Nigeria from 2017 onwards. Our samples are predominantly collected from the South South and South East regions, and do not include the period closer to the estimated emergence or the start of the epidemic in 2017. However, as Southern regions represented the highest number of cases throughout the epidemic from 2017 to the 2022 resurgence, it is unlikely that the geographic distribution of samples represents a strong sampling bias.

Controlling the ongoing mpox epidemic in Nigeria is impeded by inequities of access to resources such as diagnostics, vaccines and therapeutics. After the WHO declared mpox a Public Health Emergency of International Concern, high income countries increased and deployed their stockpiles of smallpox vaccines, including third generation vaccines.^26,27^ However, these resources have not been made available to nations in Africa, despite the historic increased incidence, morbidity and mortality of the disease on the continent.^28–30^ Tecovirimat, a smallpox antiviral effective against mpox, has also not been made accessible for African nations.^28^ Without access to therapeutics and vaccines, transmission cannot be reduced in either the sustained human epidemic or in populations at high risk for recurrent spillovers from the reservoir. Ongoing zoonotic and human transmission in Africa does not only increase the probability of re-emergence, future global epidemics and potential viral adaptation but, vitally, results in preventable morbidity and mortality in endemic countries. The global community can no longer afford to neglect mpox in Africa or perpetuate inequities in therapeutic access in our vulnerably connected world.

## Methods

### Ethics declaration

No ethical approval was required for this study as it is based on data from Nigeria’s national surveillance program, collected by the Nigeria Centre for Disease Control. Under the program, individual written or oral informed consent was obtained from all suspected mpox cases. Informed consent for children was obtained from their parents or recognized guardians.

### Sampling

Samples were collected by laboratory personnel and Local Government Area (LGA) Disease Surveillance and Notification Officers (DSNOs) equipped with appropriate personal protective equipment (PPE), adhering to the guidelines outlined in the Nigeria Centre for Disease Control and Prevention (NCDC) National Monkeypox Public Health Response Guidelines.^31^ Samples comprised: swabs from the exudate of vesicular or pustular lesions, lesion crusts obtained during the acute rash phase, whole blood collected in ethylenediaminetetraacetic acid (EDTA) or plain/non-anticoagulated tubes. All samples were labelled with case information and stored at 2-8°C during transport to either the NCDC National Reference Laboratory (Gaduwa-Abuja) or the Central Public Health Laboratory (Yaba-Lagos). On arrival, the crusts and swabs were eluted, while the serum/plasma was separated from the red blood cells. Subsequently, these components were stored at ultralow temperatures of ≤-70°C at the NCDC biorepository.

### Genome Sequencing

Enrichment bead-linked transposomes was used to tagment the extracted DNA and enriched using the Illumina-rna-prep enrichment with the VSP panel. Libraries were quantified using dsDNA BR Assay, normalized to a concentration of 0.6nM and sequenced on the Illumina NovaSeq 6000 platform with a read length of 151 base pair paired end at the African Centre of Excellence for Genomics of Infectious Diseases (ACEGID), based at Redeemer’s University, Ede, Nigeria.

### Genome Assembly

We performed initial *de novo* assembly with the viral-ngs pipeline, followed by reference based assembly with an in-house pipeline.^32^ Briefly, we mapped reads against a Clade IIb reference genome (NC_063383, an early hMPXV-1 genome from Nigeria) with *bwa-mem*^33^, and called consensus using samtools^34^ and iVar.^35^

### Genomic dataset curation

We combined our 112 genomes with all high-quality, publicly available Clade IIb MPXV genomes from Genbank (as of August 2023). This included all Clade IIb genomes from 2017-2022 sampled in Nigeria, sequences from non-endemic countries with a travel history to Nigeria and two older sequences sampled in Nigeria in 1971 and 1978 (accession numbers KJ642617 and KJ642615, respectively). We included a single representative of the global outbreak lineage B.1, as it was not our primary focus. In total, our dataset consists of 202 sequences.

### Phylogenetic Analysis

We aligned our dataset to the Clade IIb reference genome (NC_063383) using the ‘squirrel’ package (https://github.com/aineniamh/squirrel) developed by O’Toole *et al*.^11^ The alignment was trimmed, and the 3’ terminal repeat region and regions of repetition or low complexity were masked.

We investigated the preliminary placement of our sequences in a phylogeny of all available mpox genome sequences from Genbank across clades. We reconstructed the complete MPXV phylogeny with IQ-TREE v2.0, under the Jukes-Cantor substitution model.^36^ We identified three B.1 lineage sequences in our dataset. To investigate whether these sequences represented re-importations to Nigeria, we reconstructed a phylogeny with 769 B.1 genomes from Genbank. We confirmed the sequences represented re-importations of B.1 into Nigeria and excluded them from subsequent analyses (data not shown).

We reconstructed a phylogeny for Clade IIb alone under the same parameters as the global phylogeny. We rooted our phylogeny to KJ642615, a sequence sampled in Nigeria in 1978, as it is notably divergent from the remaining Clade IIb diversity. We collapsed all zero branch lengths. We performed ancestral state reconstruction with IQ-TREE2 across the Clade IIb phylogeny.^36^ We mapped all nucleotide mutations that occurred across the phylogeny to internal branches using tree traversal, excluding missing data. Additionally, we catalogued the dimer genomic context of all C→T or G→A mutations, as described by O’Toole *et al*.^11^ We classified our sequences into lineages under the nomenclature developed in Happi *et al.*^13^ using Nextclade.^37^

### Modelling APOBEC3-mediated evolution

We adopted a similar approach described by O’Toole *et al*.^11^ to analyse the evolutionary dynamics of hMPXV-1 in the software package BEAST^38^ with the BEAGLE high-performance computing library.^39^ First, we partitioned the Clade IIb alignment into two distinct partitions. The first partition comprised sites with potential APOBEC3 modifications (specifically C→T and G→A substitutions in the dinucleotide context TC and GA), along with target sites (e.g. C and G) that were conserved. In this partition, we masked all other sites as ambiguous nucleotides. This partition represents APOBEC3 mutations relative to the target APOBEC3 sites. The second partition inversely contained sites with the APOBEC3 target sites masked. The APOBEC3 alignment comprised 24 680 unmasked sites, whereas the non-APOBEC3 alignment comprised 172 529 sites. We used the standard nucleotide GTR+G substitution model with four distinct rate categories for the non-APOBEC3 partition.

For the APOBEC3 partition, we developed a substitution process where we categorise the nucleotides as modified (T) and unmodified (C). We used a two-state continuous-time Markov chain with an asymmetric rate to permit C→T mutations but not the reverse.

We used a two-epoch model to estimate the time of MPXV’s emergence in the human population. Under this model, the evolutionary rate transitions from the background rate (i.e. non-APOBEC3 rate, driven by polymerase error rate) to the APOBEC3 rate at a specific time point *tp* for the APOBEC3 partition. We parameterized this transition time as tp = tMRCA(Lineage A) + x, where x is a free parameter in BEAST representing the pre-sampled transmission history before the most recent common ancestor (MRCA) of sampled Lineage A viruses.^11^ We incorporated a local clock to scale the mutation proportion attributed to APOBEC3 activity across the branches up to the transition time. We allowed the non-APOBEC3 partition to evolve under the background evolutionary rate across the entire phylogeny.

We additionally used a two-phase coalescent model: the tree from the MRCA (Lineage A) onward was modelled with an exponential growth model, with the earlier phase modelled as a constant-population size coalescent model. We estimated the virus’ doubling time relative to the transition time. The doubling time is expressed as *log*(*2*)/*growth rate*, with the growth rate estimated from the exponential model. We further investigated Lineage A’s epidemic growth pattern using a non-parametric coalescent Skygrid model with 12 change points over 10 years, excluding the zoonotic portion of the tree. For each model, we ran two independent chains of 100 million states to ensure convergence, discarding the initial 10% of each chain as burn-in. The chains were then combined with LogCombiner. For all subsequent analyses, we assessed convergence using Tracer, and constructed a maximum clade credibility (MCC) tree in TreeAnnotator 1.10.^40^

### Geographic history of hMPXV-1 in Nigeria

#### Discrete phylogeographic analysis

To investigate the spread of hMPXV-1 across Nigeria, we reconstructed the timing and pattern of geographic transitions across Nigerian states under an asymmetric discrete trait analyses.^41^ We used Bayesian stochastic search variable selection (BSSVS) to infer non-zero migration rates and identify statistically supported migration routes. We used a non-parametric skygrid coalescent tree prior, with 12 change points distributed over 10 years as described above.^42^ We combined two independent MCMC runs of 50 million states each, sampling every 2000 states and discarding the respective initial 10% of trees as burn-in. We confirmed all ESS values are above 200.

We used a Markov jump counting procedure to investigate the timing and origin of geographic transitions, or Markov jumps, across the full posterior to account for uncertainty in phylogeographic reconstruction.^43^ We used the TreeMarkovJumpHistoryAnalyzer from the pre-release version of BEAST 1.10.5 to obtain the Markov jumps from posterior tree distributions.^19^ Using the tree distribution annotated with Markov jumps, we performed the persistence analysis on a month-to-month interval to calculate the percentage of lineages that persisted in their ancestral state for each Nigerian state and region.^19^ We used the PersistenceSummarizer from the pre-release version of BEAST 1.10.5.

We also performed all phylogeographic analyses on a regional level, to account for the uneven distribution of sequences across Nigerian states. We categorised the states into the six geopolitical zones of Nigeria.^44^ We have a limited number of sequences from Northern Nigeria, which had very low epidemic incidence from 2017 - 2022. To account for this, we combined the North West and North East zones into a single North category. All trees were visualised using baltic (https://github.com/evogytis/baltic).

#### Continuous phylogeographic analysis

We performed a continuous phylogeographic analysis to quantify the dispersal of hMPXV-1 across Nigerian states. We assigned each sequence a latitude and longitude that matched the local government area or village of collection. We used the two-epoch and skygrid coalescent model described above, with a Cauchy distribution to model the among-branch heterogeneity in dispersal velocity.^45^ We ran two independent MCMC chains of 50 million states, sampling every 2000 states. We combined the chains after discarding 10% of the states as burn-in.

#### Discrete phylogeographic analysis using a sparse General Linear Model (GLM)

To investigate the drivers of the transmission dynamics of the hMPXV-1 epidemic, we used a sparse generalised linear model (GLM) in a continuous-time Markov chain (CTMC) diffusion framework.^46^ We considered 19 covariates in the model, including epidemiological case counts relative to the numbers of sampled genomes per locations, demographic data, and geographic and economic factors (Extended Data Table 1). Besides the location-specific predictors, which were encoded as binary variables to reflect migration patterns, we log-transformed and standardised all other predictors. This standardisation involved adding a pseudo-count to each entry to ensure a robust analysis.^19^ We ran two independent MCMC chains of 50 million iterations, sampling every 2000 iterations. We combined the resulting posterior distributions after removing the initial 10% as burn-in.

### Covariate collation

Covariates (Extended Data table 1) included in the GLM were collected from the following sources: Epidemiological data was obtained from the Nigerian CDC; Economic covariates were sourced from Okeowo el al.^47^; Population covariates were sourced from the Bulletin of the National Bureau of Statistics^443^; HIV prevalence was obtained from the PEPFAR program^48^; Drug use statistics were obtained from the National Bureau of Statistics.^49^ The distance by road travel between each states was calculated from Google Maps.

### Geographical metadata

Administrative level 2 (admin2) metadata for the sampling location of sequences in the dataset were mapped to official admin2 as found in the Global Administrative Database (GADM, https://gadm.org).

### Delphy

We are preparing a separate, detailed paper on Delphy.^22^ Here, we present only the essential details to motivate the public health surveillance application we describe in the main text. Delphy is a reframing of Bayesian phylogenetics in terms of Explicit Mutation-Annotated Trees (EMATs), which exploits the near-parsimonious nature of trees over genomic epidemiology datasets with limited genetic diversity to gain significant efficiencies. See Supplementary Information for a more precise formulation.

In the version of Delphy adapted for mpox, we classify each site as having APOBEC3 context according to the sequence of the first sample in the input file (with missing sites replaced by the consensus across all input sequences). A site has APOBEC3 context if it and the preceding site form a T**C** or a T**T** dimer, or if it and its subsequent site form a **G**A or **A**A dimer. For sites without APOBEC3 context, we use a transition rate matrix for a Jukes-Cantor evolutionary model with mutation rate *μ* and no site-rate heterogeneity:

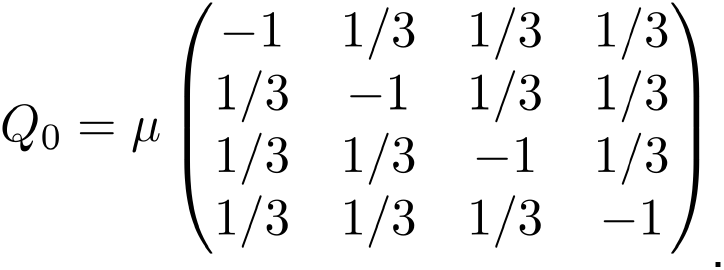

In sites with APOBEC3 context, we add transitions from C->T and G->A at a rate *μ*\*, as follows:

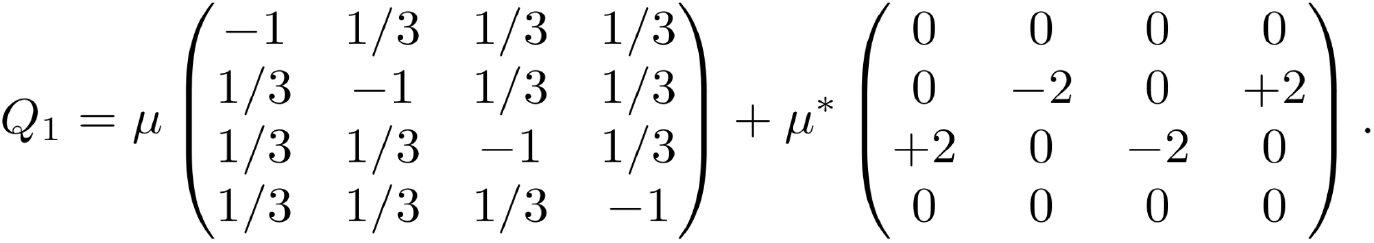

The factor of 2 is chosen to match the conventions used in the BEAST runs to define the APOBEC3 rate (the effective rate at which transitions would be observed in a sequence of APOBEC3 sites that are 50% C and 50% T).

The above setup retains the essence of that of the BEAST runs but differs minimally in its details. By deciding on the APOBEC context using the first sequence, we avoid having to previously estimate an ML tree and extract its root sequence (the assumption is that the APOBEC3 context of the vast majority sites is stable throughout the entire tree), and allows us to handle arbitrary new datasets, possibly aligned to a different reference, without complications. By preserving the polymerase mutation mechanism in sites with APOBEC3 context (i.e., the first term in the definition of Q_1_), we avoid having to identify only the sites that have not yet mutated (i.e., only the T**C** and **G**A sites) and avoid technical difficulties with completely irreversible transition matrices. For simplicity, we also use a simple Jukes-Cantor model with no site rate heterogeneity instead of a more sophisticated GTR model with 4 gamma rate categories. Delphy does not yet implement tip-date sampling, so tips with uncertain dates are imputed to the middle of the date range. Since Delphy does not yet implement a Skygrid population model, we model the viral population as growing strictly exponentially (see Figure∼2B). Finally, by design, we have not implemented a transition between a pre-spillover and post-spillover portion of the tree, as our interest is in public health response to a growing outbreak that is known to be spreading through humans. We suspect this is the dominant factor in the discrepancies between the tMRCAs and mutation rates of the main BEAST run in Figures 2 and 3 and the Delphy runs. The long branches near the top of the tree extending to the late 19th century likely exhibit purifying selection that reduces their observed substitution rate^17^, which depresses the estimate of the polymerase-driven mutation rate and pushes tMRCAs towards the past.

## Data availability

All sequences are available on Genbank under Accession numbers PP852943 - PP853055. All other data are available at https://github.com/andersen-lab/hMPXV-1_Nigeria, or upon request.

## Code availability

All code to run the analyses is available in https://github.com/andersen-lab/hMPXV-1_Nigeria. Delphy’s code is available at https://github.com/broadinstitute/delphy and https://github.com/fathominfo/delphy-web.

## Acknowledgements

This work is made possible by support from Flu Lab and a cohort of generous donors through TED’s Audacious Project, including the ELMA Foundation, MacKenzie Scott, the Skoll Foundation, and Open Philanthropy. This work was supported by grants from the National Institute of Allergy and Infectious Diseases grants U01HG007480 (H3Africa), U54HG007480 (H3Africa), U01AI151812 (WARN-ID), U19AI135995 (CViSB), U19AI110818 (GCID), R01AI153044 and R01AI162611.). This work was also supported by the World Bank grants projects ACE-019 and ACE-IMPACT; The Rockefeller Foundation (Grant #2021 HTH); The Africa CDC through the African Society of Laboratory Medicine [ASLM] (Grant #INV018978), and the Science for Africa Foundation. Ifeanyi Omah is supported by the Wellcome Trust Hosts, Pathogens & Global Health program [Wellcome Trust, Grant number 218471/Z/19/Z] in partnership with Tackling infectious Disease to Benefit Africa, TIBA. PL is supported by the European Union’s Horizon 2020 project MOOD (grant agreement no. 874850) and by the Research Foundation - Flanders (‘Fonds voor Wetenschappelijk Onderzoek - Vlaanderen’, G0D5117N, G005323N and G051322N). We thank Advanced Micro Devices, Inc. for the donation of massively parallel computing hardware.

## Author Contributions

C.H., P.S., R.N., I.J., I.A., conceptualized the study. E.P., I.F.O., P.V., A.M, A.O.T., P.L., M.A.S., K.G.A., A.R., C.H. contributed the methodology. P.V., K.G., A.O.T., P.L., M.A.S., A.R. provided software. E.P., I.F.O., P.V., A.M. performed formal analysis. E.P., I.F.O., P.V., A.M., A.O.A, A.E.S, M.I.A., O.O.E, O.A.O., A.O., P.E., O.A., A.E., O.E., C.C, K.S., A.Akinpelu, A.Ahmad, K.I.I., D.D.D., L.L.M.E, M.H.M.Y, F.M.F.C, M.Z., A.R. conducted the investigation. A.O.A, A.E.S, M.I.A., O.O.E, O.A.O, A.O., P.E., O.A., A.E., O.E., C.C, K.S., A.Akinpelu, A.Ahmad, K.I.I., D.D.D., L.L.M.E, M.H.M.Y, F.M.F.C, D.J.P, P.L., G.M., S.K.T., Y.K.T., O.F., A.H., M.A.S., K.G.A., A.R., R.N., C.I., I.J., I.A., provided resources. E.P., I.F.O., C.T.T, J.R.O., A.R., K.S. curated data. E.P., I.F.O., P.V. wrote the original draft of the manuscript. All authors reviewed and edited the manuscript. E.P., I.F.O., P.V. performed visualization. C.H., R.N., C.I., I.J., I.A., P.S., A.R., K.G.A supervised the study. E.P., O.F., C.H., A.E. undertook project administration. C.H., P.S., I.A., K.G.A. acquired funding.

## Extended Data

**Extended Data Figure 1:**
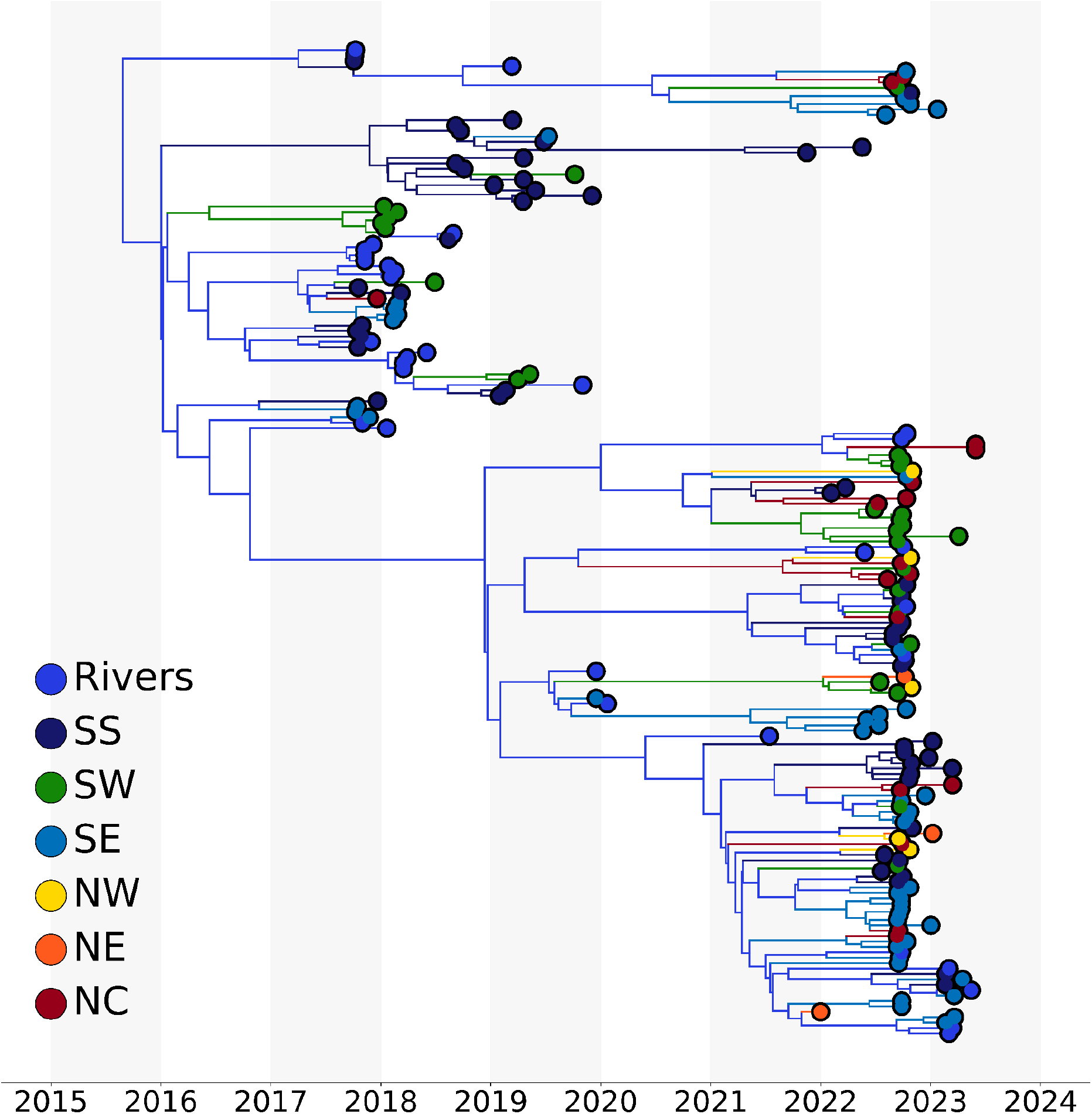
Phylogeographic analyses of Clade IIb in Nigeria. The branches of the MCC are coloured by source state, as per legend. Non-Rivers state were grouped by region. SS: South South; SW: South West; SE: South East; NW: North West; NE: North East’ NC: North Central.

**Extended Data Figure 2:**
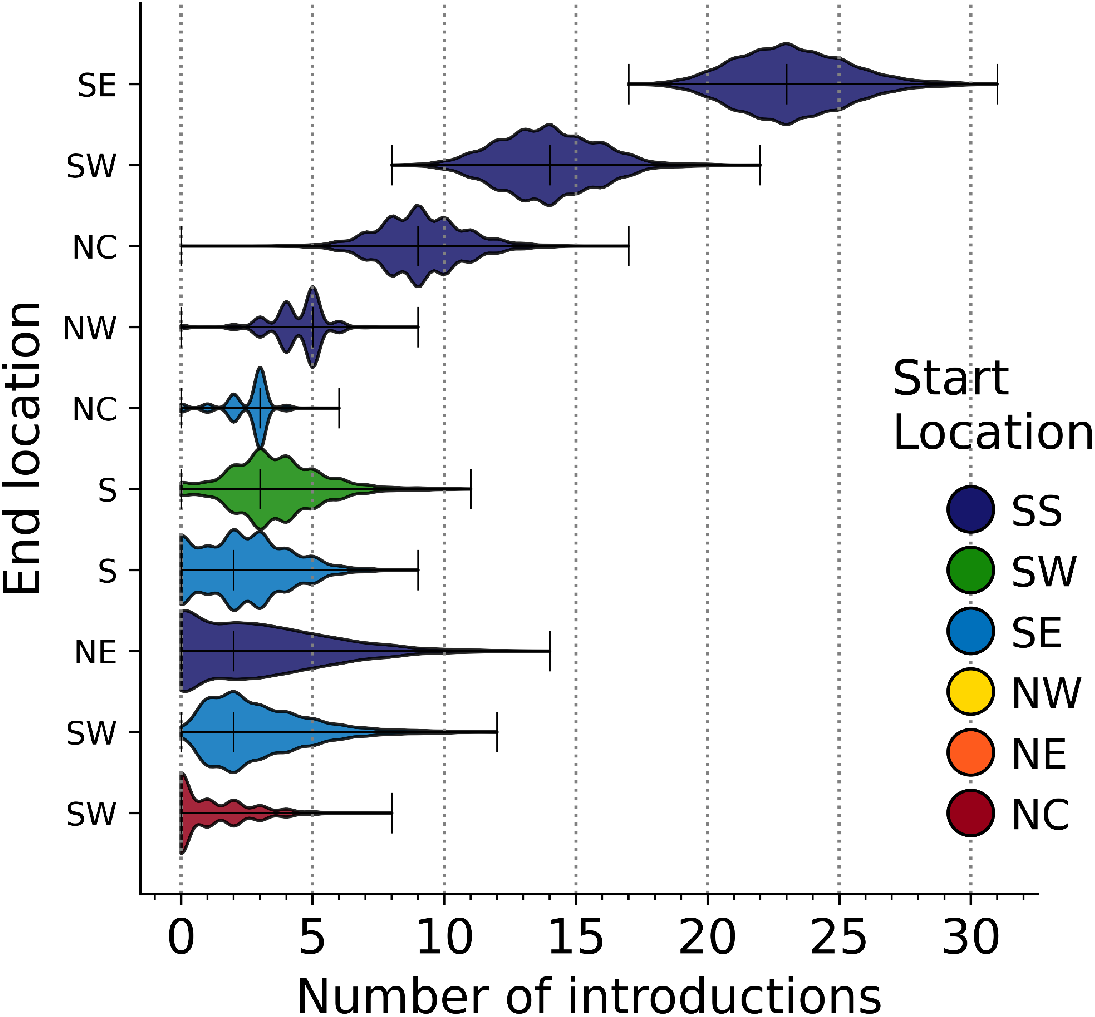
Total number of introductions by region from each start region, annotated as per legend in colour, to each end location on the y-axis.

**Extended Data Figure 3:**
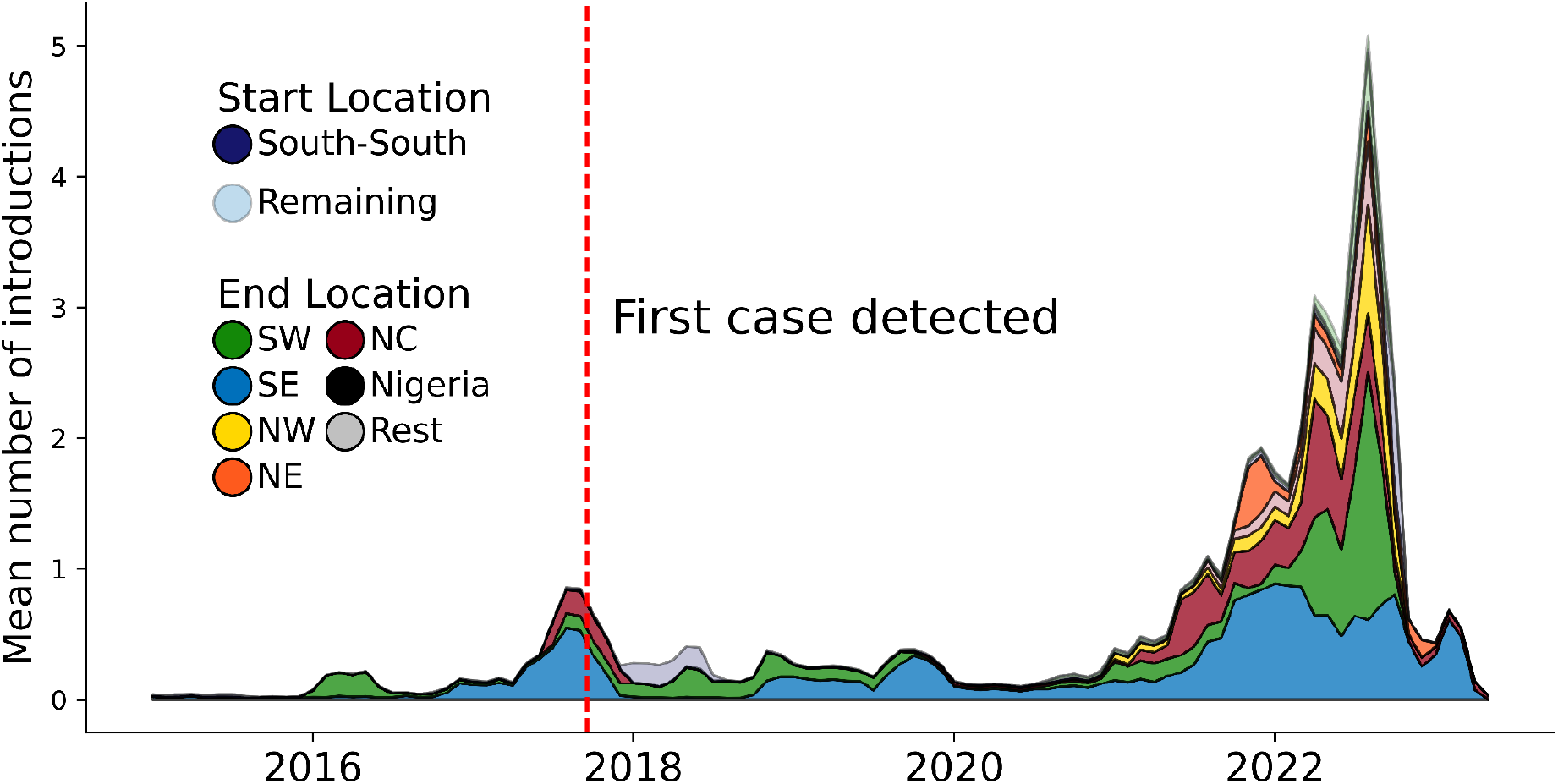
The distribution of the number of introductions across time by region. The end location state is coloured by region, as per legend. The start location is highlighted by transparency: all introductions originating from the South-South region are presented with no transparency, whereas introductions originating from other regions are transparent.

**Extended Data Figure 4:**
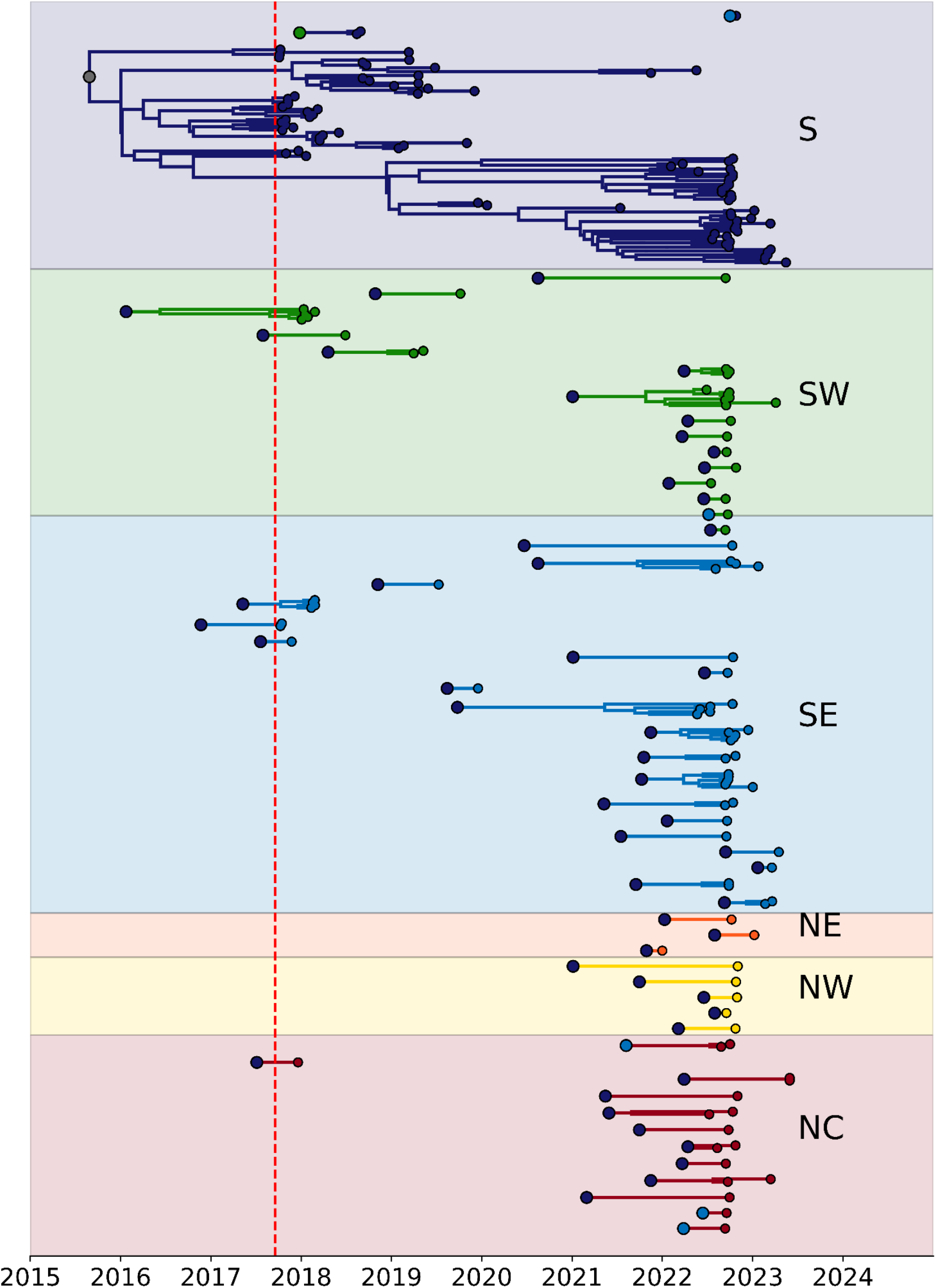
Persistence of transmission chains across all regions, as annotated in text. The start of each transmission chain is coloured by its region of origin. The red line indicates the date of report for the first case in Bayelsa.

**Extended Data Figure 5:**
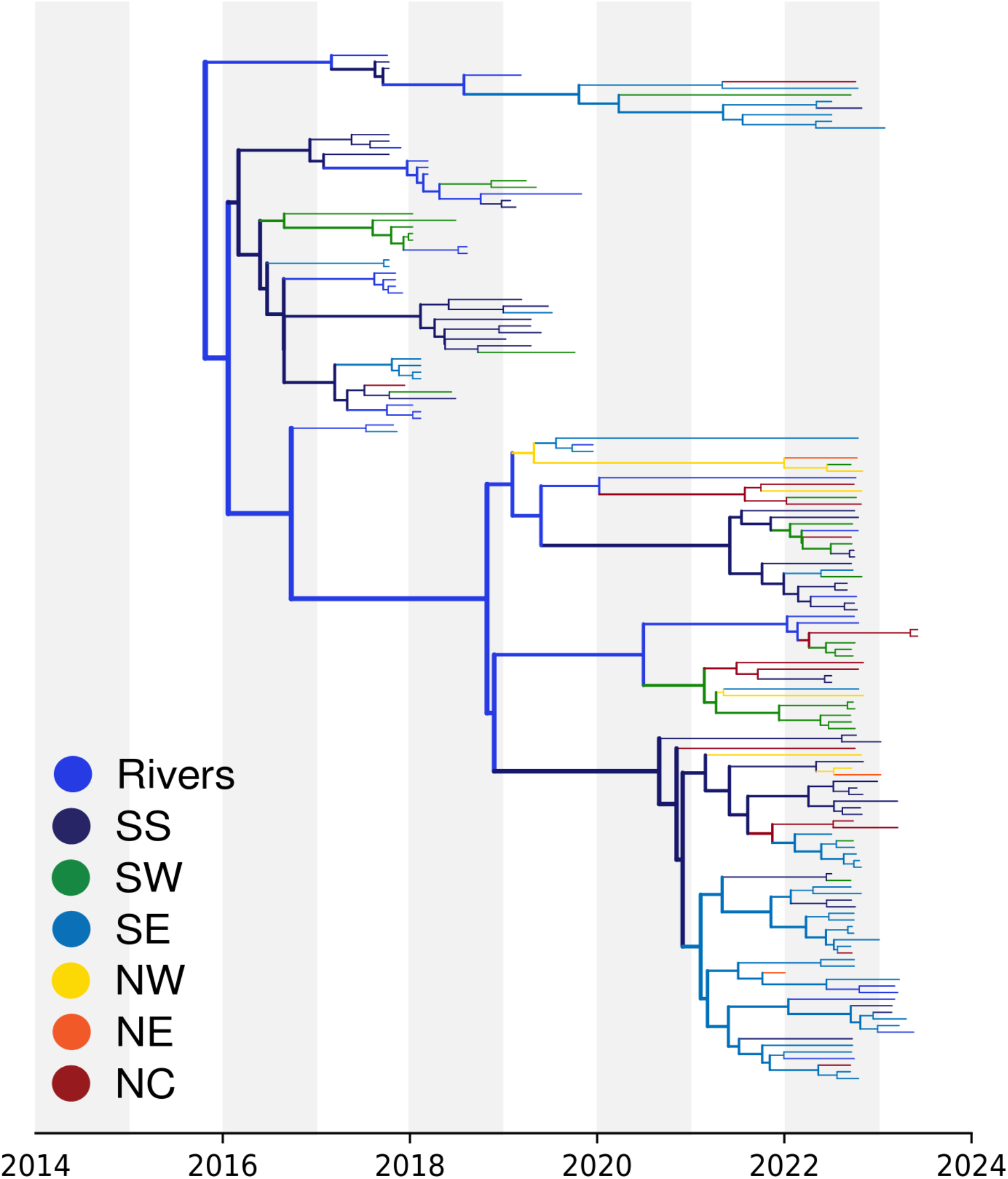
Delphy analog to Extended Data Figure 1. Although Delphy’s naive parsimony propagation of geographical labels on the MCC is particularly crude, the basic outline of the geographical spread is clearly recovered.

**Extended Data Table 1:** Covariates for GLM analyses.

| Covariates Type | Abbreviation | Covariates description | 863 |
| --- | --- | --- | --- |
| Administrative | Lagos(O) | Lagos as origin of infection | 864 |
| Administrative | Lagos(D) | Lagos as Destination of infection | 865 |
| Administrative | Rivers(O) | Rivers as origin of infection | 866 |
| Administrative | Rivers(D) | Rivers as Destination of infection | 867 |
| Demographic | Pop Size(O) | Origin population size, log-transformed, standardised | 868 |
| Demographic | Pop Size(D) | Destination population size, log-transformed, standardised |  |
| Demographic | Pop Dens(O) | Origin population density, log-transformed, standardised |  |
| Demographic | Pop Dens(D) | Destination population density, log-transformed, standardised |  |
| Demographic | Time100k (O) | Average log-transformed travel time in minutes to the closest city with a population of over 100,000 from the origin location, standardised. |  |
| Demographic | Time 100k (D) | Average log-transformed travel time in minutes to the closest city with a population of over 100,000 from the destination location, standardised. |  |
| Economics | GDP | Economic output, log-transformed, standardised |  |
| Epidemiology | Residuals(O) | Sampled genomes relative to the incidence case in Origin log-transformed, standardise |  |
| Epidemiology | Residuals(D) | Sampled genomes relative to the incidence case in Destination log-transformed, standardise |  |
| Epidemiology | HIV(O) | HIV cases at Origin log-transformed, standardise |  |
| Epidemiology | HIV(D) | HIV cases at Destination log-transformed, standardise |  |
| Epidemiology | Drug use(O) | Drug use at the origin log-transformed, standardise |  |
| Epidemiology | Drug use(D) | Destination Drug use, log-transformed, standardise |  |
| Geography | GCDistances | Great circle distances between the locations' population centroids, standardise log-transformed, |  |
| Geography | RDistances | Road distance by driving, log-transformed, standardise |  |

