## Supplementary_Information for "Genomic epidemiology uncovers the timing and origin of the emergence of mpox in humans"

### Overview of Delphy

While we are preparing a separate, detailed manuscript on Delphy [1], the tool is already available at `https://delphy.fathom.info`, with source code at `https://github.com/broadinstitute/delphy` and `https://github.com/fathominfo/delphy-web`. Here, we present only the essential idea.

Delphy models phylogenies using Explicit Mutation-Annotated Trees (EMATs), which are an extension of the Mutation-Annotated Trees (MATs) introduced by UShER [2]. An EMAT is a MAT where every node and every mutation along a branch occurs at a specific time. Since mutations are explicit, every point $x$ on the tree has a specific sequence.

The posterior distribution over EMATs is

$$P(\mathcal{T}, \boldsymbol{\theta}) \propto \left\{ \prod_{\ell \in \xi(r)} \pi^{(\ell)}_{s^{(\ell)}_r} \right\} \cdot \exp\left[ -\int_{x \in \mathcal{T}} \lambda(x)\, dx \right] \cdot \prod_{(a,\ell,b) \in \mathcal{M}} Q^{(\ell)}_{ab} \cdot \pi(\mathcal{T}, \boldsymbol{\theta}). \tag{1}$$

The notation is as follows:

- $Q^{(\ell)}_{ab}$ is the element of the transition rate matrix for site $\ell$ that encodes the rate at which a site with state $a$ mutates to a state $b$;
- $Q^{(\ell)}_a := -Q^{(\ell)}_{aa}$ is the magnitude of one of the diagonal elements of this matrix;
- $\lambda(x) := \sum_{\ell \in \xi(x)} Q^{(\ell)}_{s^{(\ell)}_x}$ is the sequence-dependent genome mutation rate at point $x$ on the tree;
- $\xi(x)$ is the set of sites for which the state at at least one tip downstream of $x$ is not missing (these are used by our scheme for dealing with missing data, *N-pruning*, which we describe in [1]);
- $s^{(\ell)}_x$ is the state of site $\ell$ in the sequence at point $x$ on the tree;
- $\pi^{(\ell)}_a$ is the stationary state distribution of the transition rate matrix $Q^{(\ell)}_{ab}$, defined as its zero left-eigenvector ($\sum_a \pi^{(\ell)}_a Q^{(\ell)}_{ab} = 0$);
- $\int_{x \in \mathcal{T}} h(x)\, dx$ is an integral of an arbitrary function $h(x)$ over every point $x$ in the tree $\mathcal{T}$, defined as the sum of branch integrals, each of which integrates over time along the branch;
- $\mathcal{M}$ is the set of all mutations on the tree, each of which is a tuple $(a, \ell, b, i, t)$ that denotes a mutation at time $t$ on the branch leading to node $i$ where the state at site $\ell$ changes from $a$ to $b$ (we suppress $i$ and $t$ above for clarity);
- $r$ is the root of the tree;
- $\boldsymbol{\theta}$ is the set of parameters that describes such things as mutation rates, population growth, etc.;

- $\pi(\mathcal{T}, \boldsymbol{\theta})$ encodes all the priors on the tree (e.g., the coalescent prior) and on the model parameters.

Suppose a correct statistical sampling of EMATs according to the above posterior distribution produces a set of EMATs $\{\mathcal{T}\}$. If we remove all mutation information information from these trees and preserve only their topologies and node times, we claim [1] that the result is statistically indistinguishable from a sampling of such mutation-blind trees according to the standard posterior [4],

$$P(\mathcal{T}, \boldsymbol{\theta}) \propto L_{\text{tree}}(\mathcal{T}|\boldsymbol{\theta})\pi(\mathcal{T}, \boldsymbol{\theta}). \tag{2}$$

Here, $L_{\text{tree}}(\mathcal{T}|\boldsymbol{\theta})$ is the usual tree likelihood calculated by Felsenstein pruning [3], assuming each tip sequence contains either fully specified states (‘`A`’, ‘`C`’, ‘`G`’, ‘`T`’) or fully ambiguous states (‘`N`’). Intuitively, this correspondence is nothing more than a rephrasing of the underlying continuous-time Markov chain for evolution in terms of Markov jumps instead of evolution matrices.

Delphy implements correct statistical sampling over EMATs using the posterior in Eq. (1), using a battery of specialized and efficient MCMC moves that we describe in [1]
